# Developing a multi-domain EHR foundation model for predicting Hepatitis B liver disease: a clinical perspective

**DOI:** 10.64898/2026.01.23.26344677

**Authors:** Caroline Weis, Filippo Grazioli, Giovanni Visonà, Adam Kania, Max F. Burg, Max Horn, Jonathan Golob, Patrick Schwab

## Abstract

Foundation models trained on patient electronic health records (EHRs) hold promise for transforming clinical care by enabling effective decision support and personalized healthcare delivery, but have been limited by a focus on intensive care objectives. Here we present a multi-domain transformer-based EHR foundation model designed to predict two liver disease outcomes in patients with Chronic Hepatitis B, an infection characterized by diverse and uncertain medical trajectories. Through case studies employing attention maps, we demonstrate that the transformer model identifies patterns similar to one-liners employed by clinical staff and depends on distinct clinical events to estimate disease progression. Our findings underscore both the utility and challenges of EHR foundation models in clinical care and the necessity to evaluate EHR-models on less-regimented diseases.

## 1 Introduction

Despite the existence of a safe and effective vaccine as well as chronic suppressive therapy, Chronic Hepatitis B (CHB) infection results in over 800 000 deaths annually and 1.5 million people are newly infected each year [19]. Liver cirrhosis and hepatocellular carcinoma (HCC), collectively referred to as C/HCC, are the leading causes of mortality among CHB patients [3]. The clinical progression among CHB patients to C/HCC is a continuous but non-linear processes with subtle and nonspecific early warning signs. Providing individualized risk assessments for developing liver diseases could enable early intervention and promote medication adherence to prevent severe outcomes. Their impact on the liver’s synthetic function typically occurs only at advanced stages, when hepatocyte function ultimately collapses. These medical characteristics hold the potential for large-scale EHR models to learn long-range dependencies and sequential symptoms in the medical trajectory.

Numerous studies have developed and evaluated pre-trained language models for clinical prediction tasks [17][20][16][4][13][21][12][7][8]. CLMBR [17] introduced a framework that learns patient representations from a large EHR database using a language objective and a Gated Recurrent Unit (GRU), which was subsequently extended [20] with a transformer-based architecture. Medical pre-trained models have also been adapted to time-to-event training objective [16] and state-space representations, including the incorporation of a Mamba block [4].

Most of the popular EHR datasets are focused on the intensive care unit (ICU) setting, such as MIMIC-III [5], MIMIC-IV [6]. ICU care is notable for intense short-term monitoring, relatively protocolized treatments and similar patient trajectories. This is in contrast to the patient trajectories from chronic diseases–with intermittent monitoring, inconsistent emergence of signs of disease progression, and variable clinical practices for monitoring. Application of EHR foundation models to indolent and chronic progressive diseases remains understudied, and it is still unclear whether predictive performance on ICU trajectories reported in the literature hold in these cases. We are among the first [13] to develop clinical pre-trained models on the Optum® de-identified Electronic Health Record data set (Optum® EHR) [2]. This is a rich multi modal dataset, including encounters, procedures, medications, clinical notes that further has de-identified longitudinal patient data.

Our contributions are as follows:

i. We develop a multi-domain EHR foundation model, and explore its utility to predict liver cirrhosis and liver cancer for chronic hepatitis B patients.
ii. We analyse attention maps of transformer blocks contained in our models and provide a medical interpretation into their decision making process in the case of insidious diseases such as C/HCC.
iii. We discuss the medical perspective on current literature evaluating EHR foundation models on diseases with stereotypical treatments paths, and argue for the necessity to evaluate on unclear patient trajectories.

## 2 Methods

We present an overview of the analysis workflow in Figure A1.

### EHR data

We trained our models on multi-center EHR data [2], which comprises de-identified, longitudinal records from over 100 million patients across more than 7,000 U.S. hospitals and clinics. The database captures multi-domain data, extracted from structured medical information and written medical notes—including patient demographics, diagnoses, and prescriptions—recorded between 2007 and 2024 (see Appendix A.1 for more details). The analysis of liver disease prediction focuses solely on a target population of patients living with chronic hepatitis B (CHB). For finetuning and an end-to-end trained transformer, we therefore define the *CHB target population* as all patients with at least one CHB diagnosis code, resulting in a dataset comprised of more than 78 000 patients (see Table A1). To obtain our pre-training dataset, we filtered the full EHR database for patients not contained in the CHB target population and for medical codes occurring in no less than 7 000 patients and included only patients with between 20 and 600 events. The resulting pretraining dataset is comprised of over 55 million patient trajectories with an average of 168 events per patient.

The tables considered in the presented analysis are *Patients* (e.g. birth year, gender, ethnicity), *Diagnosis* (e.g. ICD9, ICD10, SNOWMED codes), *Labs* (e.g. LOINC codes), *Immunization* (vaccinations), *Observation* (numerical e.g. SBP, temperature), *Prescription* (e.g. NDC codes), *Visit* (categorical visit types e.g. inpatient or emergency), *Procedures* (e.g. CPT4, ICD9, SNOMED codes), and *Administrations* (e.g. NDC codes).

The EHR data includes in-patient stays with intensive surveillance, yielding up to 26 500 recorded events per patient. To reduce the sequence length and the dominance of high-surveillance periods, events are aggregated over monthly intervals. This aggregation window corresponds to the medically relevant period for detecting changes in liver health. Let *T*_max_ be the maximum number of months with at least one event recorded for all patients in a batch, then the batch can be represented by a tuple of patients *P*_*i*_, with each *P*_*i*_:

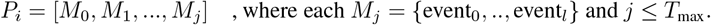

Given the expanding literature on clinical pre-trained models, the Medical Event Data Standard (MEDS)[1] has been proposed to maximize interoperability across datasets, tools, and model architectures. Our work follows the MEDS standard.

### Endpoint definition

We predict the probability of developing cirrhosis or liver cancer after four different time delays (1, 3, 5, and 10 years). These time delays correspond to clinically relevant periods for liver function deterioration. Both outcomes are studied in separate models.

### EHR tabular representation

While literature on EHR foundation models has demonstrated notable improvements over simpler baselines, historical evidence indicates that clinical machine learning models have frequently been outperformed by less complex approaches. Consequently, we challenge the EHR foundation model approach by establishing a robust and straightforward baseline using XGBoost, by converting each patient sequence into a tabular representation of the sequence. The construction of aggregated timepoints and corresponding labels match the process of the embedding models. However, instead of embedding each event for a patient, a table is constructed. Given *N*_events_ is the maximum number of event types present in the data, the tabular data representation takes the form 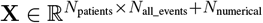. The presence of categorical features in a patient’s sequence is one-hot encoded. Numerical events are described by two columns; the first recording the numerical value (in case multiple numerical events are recorded for the same event type, the most recent measurement is taken) and the second indicating presence/absence of the value, to be able to differentiate between true zero-valued features and not recorded features. We employed `sklearn`’s [10] `SelectKBest` feature selection on the validation set with the criterion `f_classif` (ANOVA F-value) to reduce the number of features in the table from *N*_all_events_ + *N*_numerical_ to 1000.

### EHR sequence embedding

Depending on the feature domain, each medical event in time-bin *M*_*j*_ is described by an obligatory categorical feature defined by a medical code or text, and by an optional numerical value. Each event is embedded according to their event type, into a common model dimension *D*. All embeddings within the monthly aggregation time window are averaged, leading to each *M*_*j*_ in *P*_*i*_ being represented by a single embedding vector **X** ∈ ℝ^*D*^. Therefore, a batch tensor with *B* padded sequences takes the shape 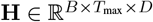.

For each *M*_*j*_, two additional embeddings are constructed and added: a trainable categorical embedding indicating age **A** ∈ ℝ^*D*^ and a fixed positional encoding **P** ∈ ℝ^*D*^. The final tensor given to the transformer **Z** is constructed as 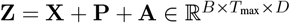.

### 2.1 An EHR foundation model

### end-to-end trained model

We base our model architecture largely on existing methods [20] [17]. The end-to-end trained model consists of a transformer encoder with *L* identical layers, employing a masked multi-head self-attention considering all subsequences for training. We combine the transformer position-wise embeddings with a binary multi-task MLP prediction head with sigmoid activation. All time horizon outcomes are treated as a multi-task prediction objective, leading to 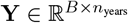. The model is trained using the `AdamW` optimiser and stopped at reaching the validation loss minimum.

#### pretrained model

For the pre-trained and fine-tuned models, an additional embedded time offset **ΔT** in days describing time passed between events is added to the input **Z**^∗^ = **Z** + **ΔT** [21]. As each position in **Z**^∗^ constitutes of multiple aggregated events, we use a multi-token prediction setup for pretraining the EHR foundation model. The pretraining prediction head is a multi-layer perceptron (MLP) providing a logit tensor for each code in the dataset 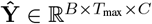, where *C* is the vocabulary size, i.e. the number of medical event codes in the dataset.

The loss function used to optimise the pretrained model consists of three parts:

1. a self-supervised multi-token objective consists in predicting the set of medical events present in the next aggregated time window, by optimising a multi-label cross-entropy loss ℒ_next_ (Eq. 1),
2. for each time window *M*_*i*_, the model fits a regression head to predict a time offset in days to the earliest event in the next window *M*_*i*+1_, using a mean squared error loss ℒ_time_ (Eq. 2),
3. and for the numerical targets in the dataset, a Gaussian prediction head estimates the mean and log-variance of the distribution through a negative log-likelihood loss ℒ_num_ (Eq. 3).

The total loss is defined as ℒ_total_ = ℒ_next_ + ℒ_time_ + ℒ_num_. Multiple model sizes were evaluated in our study (2.1.1). Results presented are based on the 170M model.

#### 2.1.1 Loss functions and training details

Given the target **Y** and the padding boolean mask 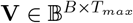, the self-supervised multi-token objective consists in predicting the set of medical events present in the next aggregated time window, by optimising a multi-label cross-entropy (CE) loss:

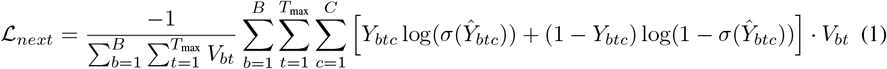

Additionally, for each token, the model fits a regression head to predict the time offset to the next time point 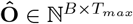 measured in days between the aggregated event tokens. The loss is a mean squared error (MSE):

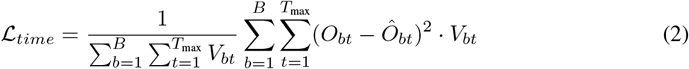

Given a numerical target **Num** 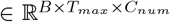 for the *C*_*num*_ numerical events of the dataset, a Gaussian prediction head estimates the mean and log-variance of the distribution. Considering a numerical mask 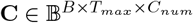 which represents if a given numerical code is included in the batch, the negative log-likelihood loss is:

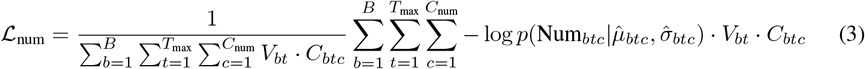

Three model sizes were evaluated in our study; 43M, 170M and 860M parameters. We obtained equally high performances for the 170M and 860M models, and are presenting the 170M model in more detail. The 43M model is trained in full precision, while mixed precision is used for the larger models. Distributed Data Parallel (DDP) training is applied to the 43M and 170M models, and DeepSpeed ZeRO-2 [11] is utilized for the 860M model. Learning rate scheduling follows a cosine annealing schedule with warm-up. Pre-trained models are fine-tuned on the CHB target population training data, initializing weights from pre-training and fine-tuning only the final transformer layer and the prediction head, with all other weights frozen.

## 3 Results

The prediction results are displayed in Table 1. The results indicate that end-to-end transformers trained directly on the CHB target population are not predicting comparatively well to the XGBoost baseline or the finetuned foundation model. While the finetuned foundation models show clear gains in performance compared to the end-to-end transformers, they reach a similar performance to but never outperform a simple tabular XGBoost baseline.

**Table 1.** Model comparison between tabular XGBoost baseline (e2e-xgb-tab), end-to-end transformer (e2e-tfm) and finetuned EHR foundation model (fm-ft-tfm). We report mean ± standard deviation over the same five fixed train-test splits used in all experiments. Metrics are area under the ROC curve (AUC) and specificity at 90 percent sensitivity (sp@90sen).

| disease | year | e2e-tfm<br>AUC | e2e-tfm<br>sp@90sen | e2e-xgb-tab<br>AUC | e2e-xgb-tab<br>sp@90sen | fm-ft-tfm<br>AUC | fm-ft-tfm<br>sp@90sen |
| --- | --- | --- | --- | --- | --- | --- | --- |
| cirrhosis | 1 | 0.74 ± 0.02 | 0.39 ± 0.03 | **0.83** ± 0.01 | 0.49 ± 0.04 | 0.80 ± 0.01 | 0.48 ± 0.03 |
| cirrhosis | 3 | 0.74 ± 0.01 | 0.39 ± 0.02 | **0.82** ± 0.01 | 0.48 ± 0.02 | 0.79 ± 0.01 | 0.48 ± 0.04 |
| cirrhosis | 5 | 0.74 ± 0.02 | 0.40 ± 0.02 | **0.81** ± 0.01 | 0.49 ± 0.03 | 0.79 ± 0.01 | 0.48 ± 0.03 |
| cirrhosis | 10 | 0.75 ± 0.02 | 0.41 ± 0.03 | **0.81** ± 0.00 | 0.49 ± 0.03 | 0.80 ± 0.01 | 0.48 ± 0.03 |
| HCC | 1 | 0.75 ± 0.03 | 0.42 ± 0.13 | **0.80** ± 0.01 | 0.41 ± 0.02 | **0.80** ± 0.02 | 0.40 ± 0.07 |
| HCC | 3 | 0.76 ± 0.04 | 0.46 ± 0.11 | **0.78** ± 0.03 | 0.40 ± 0.06 | 0.76 ± 0.02 | 0.36 ± 0.09 |
| HCC | 5 | 0.76 ± 0.06 | 0.43 ± 0.14 | **0.77** ± 0.03 | 0.39 ± 0.06 | 0.76 ± 0.03 | 0.37 ± 0.08 |
| HCC | 10 | 0.75 ± 0.05 | 0.36 ± 0.14 | **0.78** ± 0.03 | 0.39 ± 0.07 | 0.76 ± 0.03 | 0.38 ± 0.08 |

With known advantages and limitations in attention interpretability [14], we gather insights into transformer model mechanics by analyzing attention weights patterns observed regularly over a large number of patients, and showcase them in two exemplary case studies in Figure 1. We regularly observe that one attention head is focusing solely on the first event, which constitutes a *context token* of fixed patient characteristics: gender, year of birth, race, ethnicity, and geographical region (Figure 1a). Remarkably, this resembles the way that doctors and nurses communicate efficiently about patients through ‘one-liners’. The one-liner starts with a patient identifier providing clinically important information and paints a picture of the patient in the reader’s mind [15], quickly sharing a baseline pretest probability for causes of the presenting medical concern. We find the emergence of this convergent behavior in our models as quite astonishing, while also noting the weaknesses of learning misleading correlations (e.g. race as a marker for socioeconomic determinants of health). A second pattern is that of attention heads placing *full attention on the latest relevant medical event*, initially the context token, potentially switching full focus to a newly occurring event in the sequence (Figure 1b). Investigating the medical trajectory of the studied patient, attention in head no. 5 was initially fixed on the context token, then fully switched to a token for a medical code indicating abnormal findings on diagnostic imaging of liver, and switching again to a token recording a hepatic function panel. The same attention head placed attention on an abnormal findings on diagnostic imaging of liver in other patients as well. This could indicate that this specific attention head has learned to focus on hepatic function for predicting HCC.

**Figure 1.**
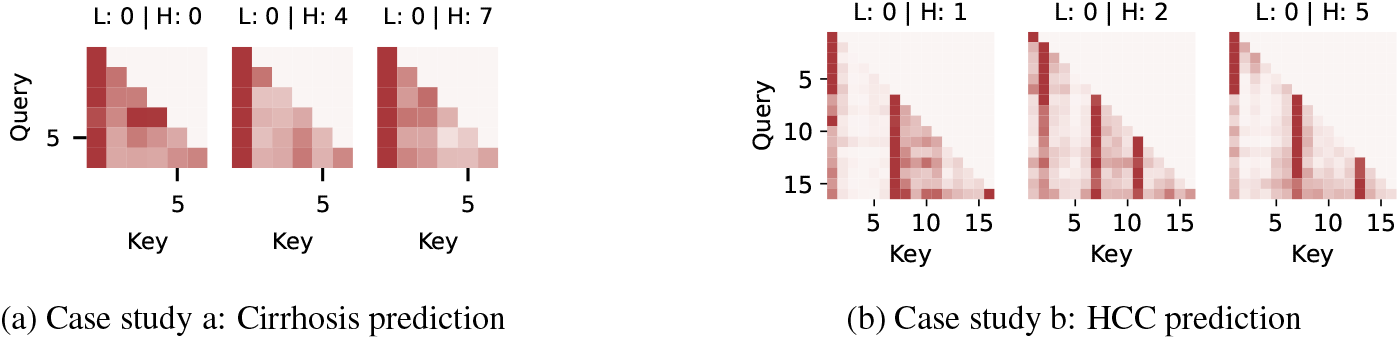
Attention weights of end-to-end transformer. Attention maps for the first model layer (L) and selected prediction heads (H) for each time index. Color scale normalized to the maximum value within each row (darkest color) and zero (brightest color). Attention across all heads and predictions for the showcased patients can be found in Figure A2.

Further, we find several indications that the EHR-models use *sequence aggregation* operations. In end-to-end transformers, performance increases with larger time aggregation windows (Figure A4). This aggregation can also be seen in the pretrained model trained on the multi-token objective, where horizontal lines in pretrained foundation models indicate that some query tokens place attention on all other key tokens, effectively leading to an averaging on the signal of all tokens (Figure A3).

## 4 Discussion

In this work we are presenting a novel multi-domain EHR foundation model intended for long-term disease monitoring, and evaluate its capability for predicting chronic progressive diseases in the case of liver cirrhosis and liver cancer in CHB patients. We have demonstrated its utility of outperforming an end-to-end transformer trained directly on the target population, while critically comparing to and demonstrating a similar performance employing a non-temporal XGBoost prediction model. Emergent behavior mirroring clinical care is observed by placing attention to the ‘one-liner’ summarizing a patient’s baseline health status. This behavior speaks to the potential of tackling difficult clinical problems. We present indications that the transformer models struggle to take advantage of the temporal aspect of the EHR sequences, and fall back to leveraging simpler techniques such as sequence aggregation to base their predictions by favoring large time aggregation windows and showing averaging steps in attention maps. We hypothesize that vanilla EHR foundation model architectures may not fully capture the complexity of medical event codes and their temporal interactions due to the relatively small number of EHR sequences compared to the training data available in language domains. The XGBoost setup likely created a more focused space of medical events by employing simple feature selection. A promising development would be to introduce greater inductive bias into the models, such as improving medical tokenization [18] or increasing training size by combining EHR datasets.

We are now further giving a medical perspective on the current literature and utilization of EHR-based foundation models: Prior work has demonstrated the utility of EHR foundation models over simpler baselines, such as logistic regression [17] and XGBoost [4]. The choice of endpoints used for performance evaluation is often motivated by the focus on ICU datasets, e.g. inpatient mortality [17] [4], long admission [17], ICU transfer [17], or readmission [17] [4]. The medical problems underlying these endpoints–e.g. heart failure, sepsis and cardiogenic shock–have well-developed management pathways that are systematically enforced by healthcare providers via payment rate benchmarks [9]. This results in most patients having a fairly regimented management pathway that tends to be documented in a stereotypical manner in health records, enabling sequence-based encoders to effectively capture and impute the continuation of these regimented medical protocols.

However, this is the exception rather than the norm; many common conditions, such as liver abnormalities, exhibit ad hoc diagnostics and less predictable treatment trajectories. As we think forward to foundational EHR-trained models as decision support tools for physicians and health systems, being able to grapple with complex and contradictory patient information offers a key opportunity for advancement in health. The presented results call into question whether the superior performance of EHR foundation models has been demonstrated in the use case of predicting patient trajectories uncertain to the medical professional. In an era of increasingly constrained provider time with patients at health check-ins, it is often these slowly progressive diseases that get missed, and only addressed when the disease has progressed to an overt, and often irreversible state. A EHR-trained model capable of flagging the subtle signs of earlier disease progression could be an invaluable partner to the primary care provider, enabling a focus on disease suppression, treatment adherence, and even direction to novel curative therapies.

## Data Availability

All data produced in the present study are available upon reasonable request to the authors

## A Technical Appendix and Supplementary Material

**Figure A1:**
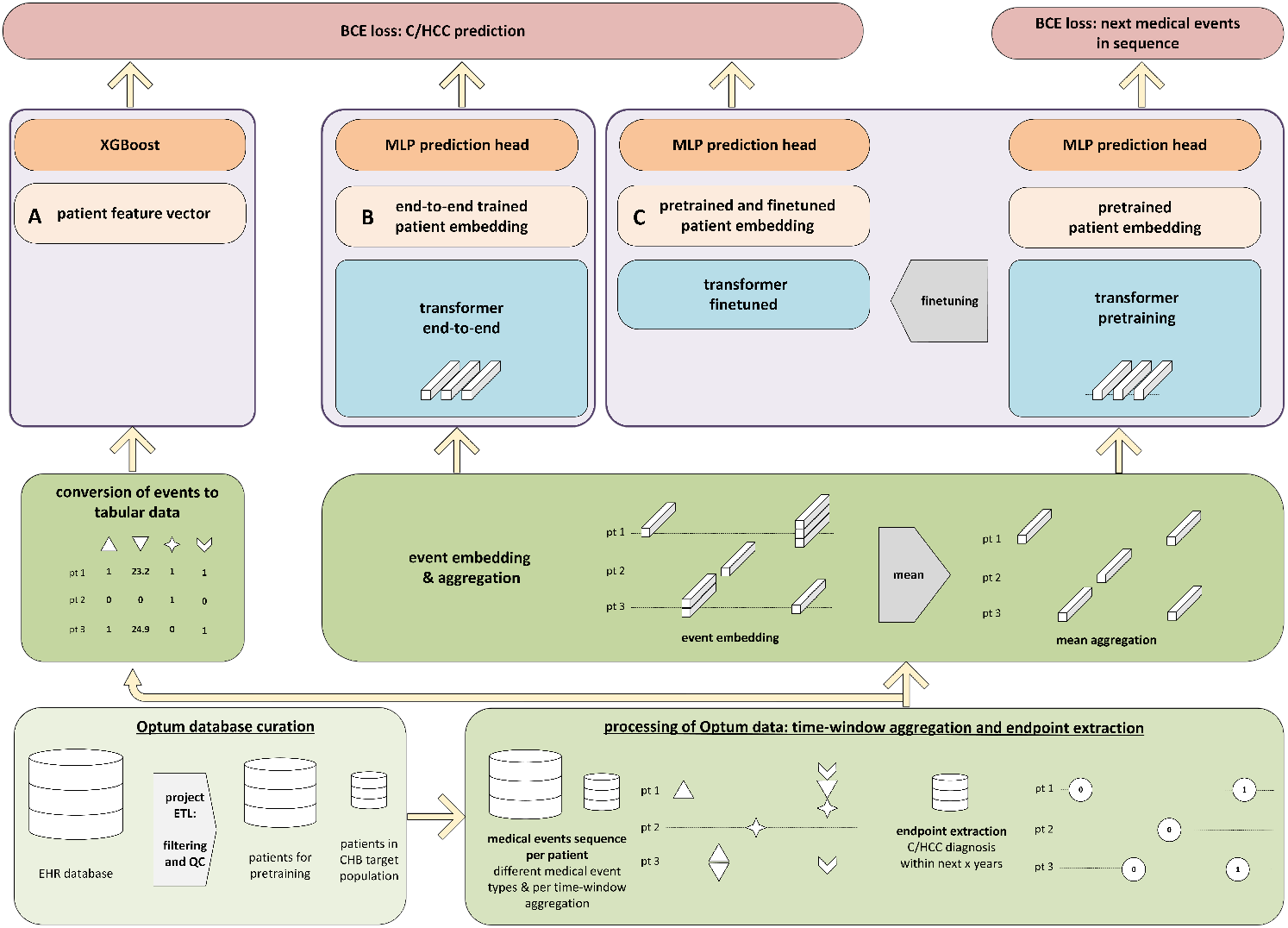
C/HCC prediction workflow overview. The workflow of the presented risk prediction models is depicted from the bottom of the plot upwards. The common initial processing includes extracting the patients with chronic hepatitis B, extracting the medical events sequence and the respective endpoints at each timepoint. The workflow then splits into three downstream paths, **A**. one using a tabular data representation defining a feature vector for each patient, **B**. directly uses a mean feature embedding to train an end-to-end transformer, and **C**. pretrains a model on a multi-token loss first and is then finetuned on the C/HCC prediction objective.

### A.1 EHR multi-domain database

Optum® de-identified Electronic Health Record data set (Optum® EHR) is a longitudinal electronic health record repository derived from dozens of healthcare provider organizations in the United States. Administrative medical data is obtained from both Inpatient and Ambulatory electronic health records (EHRs), practice management systems, and other internal systems and is processed, normalized, and standardized across the continuum of care from both acute inpatient stays and outpatient visits. The data is statistically de-identified under the HIPAA Privacy Rule’s Expert Determination method and managed according to Optum® customer data use agreements.

**Table A1:** Filter conditions for chronic Hepatitis B patients by medical code. All individuals that have at least one entry corresponding to these codes are included in the analysis. Abbr.: ‘Chronic viral hepatitis B’ was abbreviated to CHB.

| data table | Version | Code | Condition description |
| --- | --- | --- | --- |
| Diagnosis | ICD10 | B18.0 | CHB with delta-agent |
| Diagnosis | ICD10 | B18.1 | CHB without delta-agent |
| Diagnosis | ICD9 | 070.23 | CHB with delta-agent |
| Diagnosis | ICD9 | 070.33 | CHB with delta-agent without hepatic coma |
| Diagnosis | ICD9 | 070.22 | CHB without delta-agent with hepatic coma |
| Diagnosis | ICD9 | 070.32 | CHB without delta-agent without hepatic coma |
| Diagnosis | SNOMED | 61977001 | Chronic type B viral hepatitis |
| Diagnosis | SNOMED | 186639003 | Chronic viral hepatitis B without delta-agent |
| Diagnosis | SNOMED | 50167007 | Chronic active type B viral hepatitis |
| Diagnosis | SNOMED | 38662009 | Chronic persistent type B viral hepatitis |
| Diagnosis | SNOMED | 235869004 | Chronic viral hepatitis B with hepatitis D |
| Diagnosis | SNOMED | 713966008 | Occult chronic type B viral hepatitis |
| Diagnosis | SNOMED | 424340000 | CHB with hepatic coma |

#### Data access statement

The source data used for the present study were licensed from the Optum® de-identified EHR database (https://www.optum.com/), with restrictions that do not allow for the data to be redistributed or made publicly available. However, for accredited researchers, the Optum® de-identified EHR database is available for licensing at Optum, Inc. Data access may require a data-sharing agreement and may incur data access fees.

#### High positive class imbalance

Very few events get a positive class label. The positive class ratio of having either cirrhosis or liver cancer within the prediction time horizons of 1, 3, 5 or 10 years changes slightly, but ranges from 3 to 7 percent. For the end-to-end and finetuned models, the option to adjust the weight of the postive class in the BCE loss for the class ratio through `pos_weight` is given as a hyperparameter.

**Figure A2:**
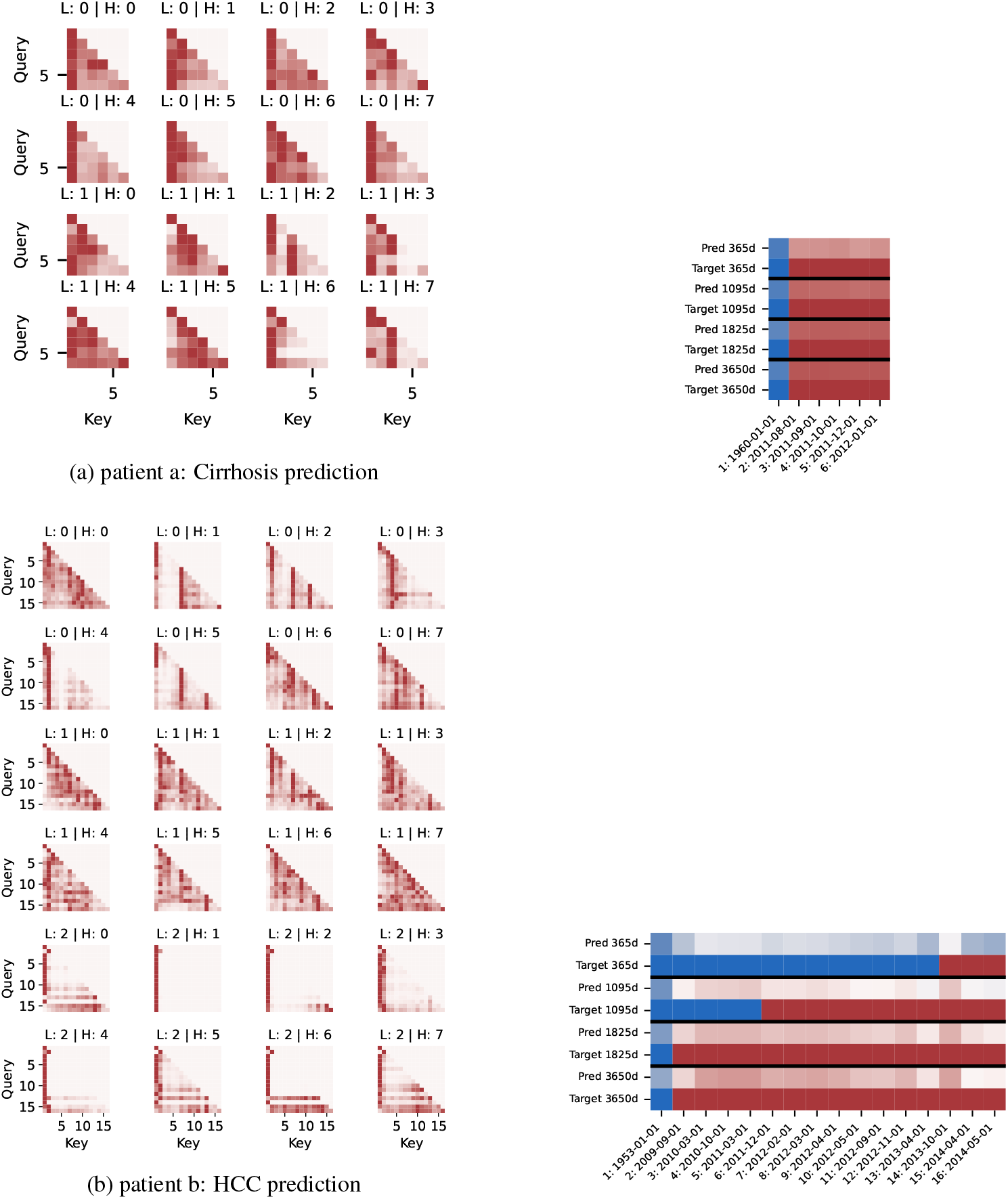
Full attention weights of end-to-end transformer. Left: Attention maps for the first model layer (L) and all prediction heads (H) for each time index. Color scale normalized to the maximum value within each row (darkest color) and zero (brightest color). Right: Predictions and labels for each time index (blue: 0, white: 0.5, red: 1).

**Figure A3:**
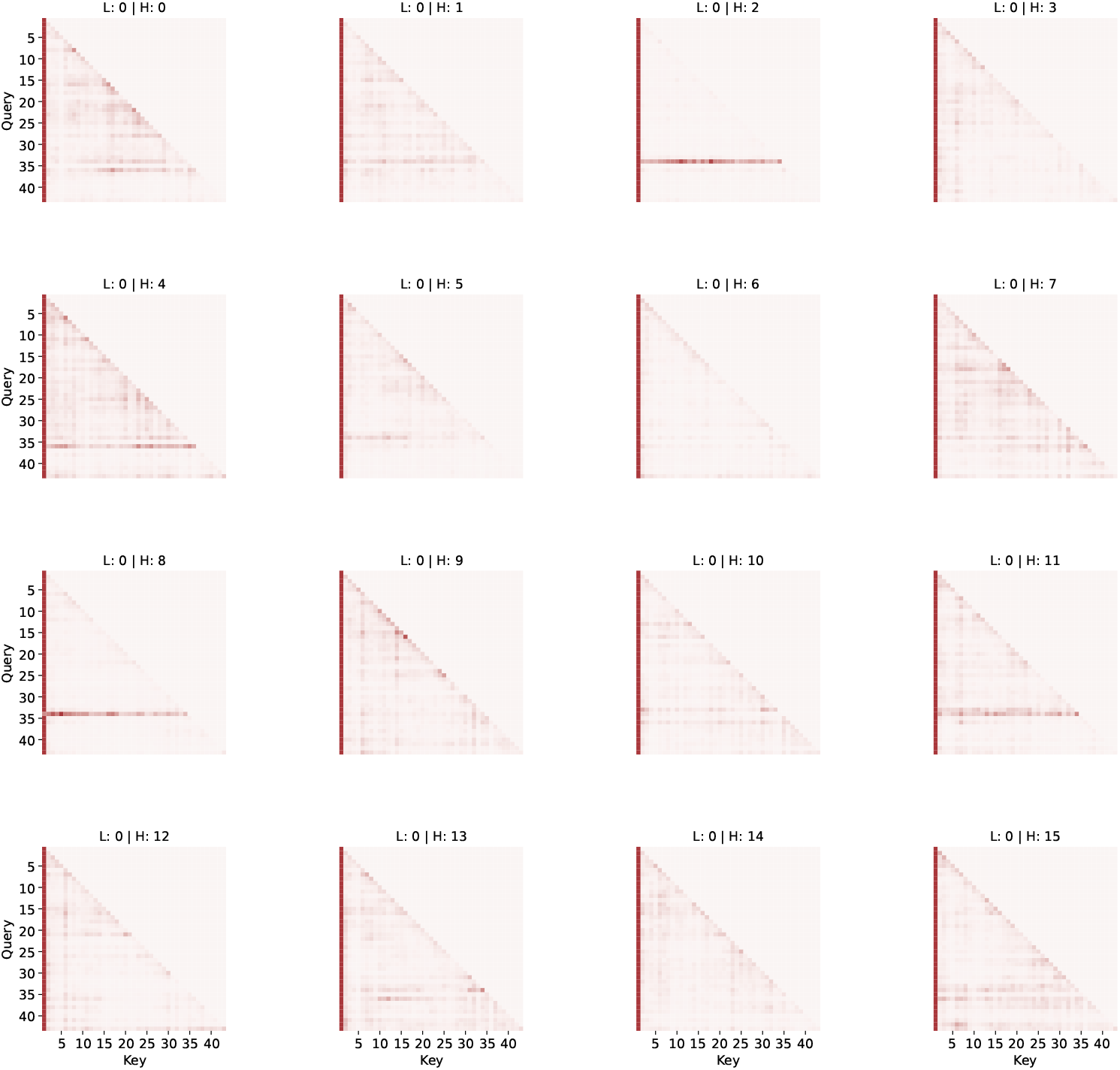
Attention maps of first layer of pretrained foundation model. High attention on the first token in the sequence can be observed. This is the context token, describing a number of set patient characteristics: gender, year of birth, race, ethnicity, and geographical region. Several heads spread out remaining attention across a wide range of tokens, giving all tokens the chance to attend to each other.

**Figure A4:**
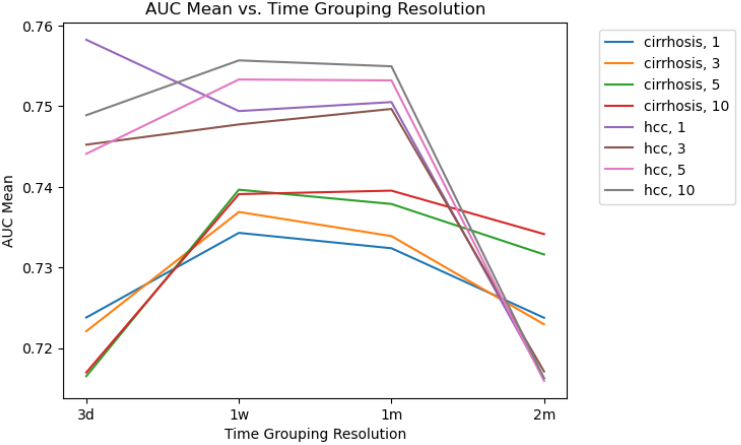
End-to-end trained transformer predictive performance vs. aggregation time window. An increase in AUC can be observed until 1 month, with a decrease for larger aggregation windows of 2 months.

## Notes

### Competing Interest Statement

C.W., F.G., G.V., A.K., M.B., J.G. and P.S. are employees and shareholders of GSK plc. M.H. is a former employee of GSK plc.

### Funding Statement

This study did not receive any funding

### Author Declarations

Optum EHR. https://business.optum.com/en/data-analytics/life-sciences/real-world-data/ehr-data.html. Optum de-identified Electronic Health Record data set (Optum EHR) is a longitudinal electronic health record repository derived from dozens of healthcare provider organizations in the United States. Administrative medical data is obtained from both Inpatient and Ambulatory electronic health records (EHRs), practice management systems, and other internal systems and is processed, normalized, and standardized across the continuum of care from both acute inpatient stays and outpatient visits. The data is statistically de-identified under the HIPAA Privacy Rules Expert Determination method and managed according to Optum customer data use agreements.

## References

[1] Medical event data standard (MEDS) format. https://github.com/Medical-Event-Data-Standard/meds.

[2] Optum® EHR. https://business.optum.com/en/data-analytics/life-sciences/real-world-data/ehr-data.html. Optum® de-identified Electronic Health Record data set (Optum® EHR) is a longitudinal electronic health record repository derived from dozens of healthcare provider organizations in the United States. Administrative medical data is obtained from both Inpatient and Ambulatory electronic health records (EHRs), practice management systems, and other internal systems and is processed, normalized, and standardized across the continuum of care from both acute inpatient stays and outpatient visits. The data is statistically de-identified under the HIPAA Privacy Rule’s Expert Determination method and managed according to Optum® customer data use agreements.

[3] D. Bixler, Y. Zhong, K. N. Ly, A. C. Moorman, P. R. Spradling, E. H. Teshale, L. B. Rupp, S. C. Gordon, J. A. Boscarino, M. A. Schmidt, Y. G. Daida, S. D. Holmberg, S. D. Holmberg, E. H. Teshale, P. R. Spradling, A. C. Moorman, J. Xing, Y. Zhong, S. C. Gordon, D. R. Nerenz, M. Lu, L. Lamerato, J. Li, L. B. Rupp, N. Akkerman, T. Zhang, S. Trudeau, Y. Zhou, K.-H. Wu, J. A. Boscarino, Z. S. Daar, R. E. Smith, Y. G. Daida, C. M. Trinacty, J. W. Lai, C. P. Wong, M. A. Schmidt, and J. L. Donald. Mortality among patients with chronic hepatitis b infection: The chronic hepatitis cohort study (checs). Clinical Infectious Diseases, 68(6):956–963, July 2018.

[4] A. Fallahpour, M. Alinoori, W. Ye, X. Cao, A. Afkanpour, and A. Krishnan. EHRMamba: Towards generalizable and scalable foundation models for electronic health records. arXiv, 2024.

[5] A. E. Johnson, T. J. Pollard, L. Shen, L.-w. H. Lehman, M. Feng, M. Ghassemi, B. Moody, P. Szolovits, L. Anthony Celi, and R. G. Mark. MIMIC-III, a freely accessible critical care database. Scientific Data, 3(1), May 2016.

[6] A. E. W. Johnson, L. Bulgarelli, L. Shen, A. Gayles, A. Shammout, S. Horng, T. J. Pollard, S. Hao, B. Moody, B. Gow, L.-w. H. Lehman, L. A. Celi, and R. G. Mark. MIMIC-IV, a freely accessible electronic health record dataset. Scientific Data, 10(1), Jan. 2023.

[7] S. J. Mataraso, C. A. Espinosa, D. Seong, S. M. Reincke, E. Berson, J. D. Reiss, Y. Kim, M. Ghanem, C.-H. Shu, T. James, Y. Tan, S. Shome, I. A. Stelzer, D. Feyaerts, R. J. Wong, G. M. Shaw, M. S. Angst, B. Gaudilliere, D. K. Stevenson, and N. Aghaeepour. A machine learning approach to leveraging electronic health records for enhanced omics analysis. Nature Machine Intelligence, 7(2):293–306, Jan. 2025.

[8] M. B. A. McDermott, B. Nestor, P. Argaw, and I. Kohane. Event stream gpt: A data pre-processing and modeling library for generative, pre-trained transformers over continuous-time sequences of complex events, 2023.

[9] Milliman. Commercial reimbursement benchmarking. https://www.milliman.com/en/insight/commercial-reimbursement-benchmarking. Accessed: October 2023.

[10] F. Pedregosa, G. Varoquaux, A. Gramfort, V. Michel, B. Thirion, O. Grisel, M. Blondel, P. Prettenhofer, R. Weiss, V. Dubourg, J. Vanderplas, A. Passos, D. Cournapeau, M. Brucher, M. Perrot, and E. Duchesnay. Scikit-learn: Machine learning in Python. Journal of Machine Learning Research, 12:2825–2830, 2011.

[11] S. Rajbhandari, J. Rasley, O. Ruwase, and Y. He. Zero: Memory optimizations toward training trillion parameter models. In SC20: International Conference for High Performance Computing, Networking, Storage and Analysis, pages 1–16. IEEE, 2020.

[12] P. Renc, Y. Jia, A. E. Samir, J. Was, Q. Li, D. W. Bates, and A. Sitek. Zero shot health trajectory prediction using transformer. npj Digital Medicine, 7(1), Sept. 2024.

[13] M. Rupp, O. Peter, and T. Pattipaka. ExBEHRT: Extended Transformer for Electronic Health Records, page 73–84. Springer Nature Switzerland, 2023.

[14] S. Serrano and N. A. Smith. Is attention interpretable? In A. Korhonen, D. Traum, and L. Màrquez, editors, Proceedings of the 57th Annual Meeting of the Association for Computational Linguistics, pages 2931–2951, Florence, Italy, July 2019. Association for Computational Linguistics.

[15] SOAPAssist. The soap note one-liner. https://soapassist.com/the-soap-note-one-liner/, 2023. Accessed: 2023-10-31.

[16] E. Steinberg, J. Fries, Y. Xu, and N. Shah. Motor: A time-to-event foundation model for structured medical records. arXiv, 2023.

[17] E. Steinberg, K. Jung, J. A. Fries, C. K. Corbin, S. R. Pfohl, and N. H. Shah. Language models are an effective patient representation learning technique for electronic health record data. arXiv, 2020.

[18] X. Su, S. Messica, Y. Huang, R. Johnson, L. Fesser, S. Gao, F. Sahneh, and M. Zitnik. Multimodal medical code tokenizer, 2025.

[19] World Health Organisation (WHO). Hepatitis B fact sheet. https://www.who.int/news-room/fact-sheets/detail/hepatitis-b.

[20] M. Wornow, R. Thapa, E. Steinberg, J. A. Fries, and N. H. Shah. EHRSHOT: An EHR benchmark for few-shot evaluation of foundation models. arXiv, 2023.

[21] Z. Yang, A. Mitra, W. Liu, D. Berlowitz, and H. Yu. TransformEHR: transformer-based encoder-decoder generative model to enhance prediction of disease outcomes using electronic health records. Nature communications, 14(1):7857, 2023.

